# *HLA-B27* and not variation in *MICA* is responsible for genotype by sex interaction in risk of Ankylosing Spondylitis

**DOI:** 10.1101/2021.12.16.21267808

**Authors:** Zhixiu Li, Allan F. McRae, Geng Wang, Jonathan J. Ellis, Tony J. Kenna, Jessica Whyte, Matthew A. Brown, David M. Evans

**Affiliations:** Centre for Genomics and Personalised Health, Queensland University of Technology, Brisbane, Australia; Queensland University of Technology (QUT), Faculty of Health, School of Biomedical Sciences, Brisbane, Queensland, Australia; Institute for Molecular Bioscience, University of Queensland, Brisbane, Australia; University of Queensland Diamantina Institute, University of Queensland, Brisbane, Australia; Centre for Immunology and Infection Control, Queensland University of Technology, Brisbane, Queensland, Australia; Department of Medical and Molecular Genetics, Faculty of Life Sciences and Medicine, King’s College London, United Kingdom; Genomics England, United Kingdom; Medical Research Council Integrative Epidemiology Unit, University of Bristol, Bristol, United Kingdom

## Abstract

Ankylosing Spondylitis (AS) is a highly heritable inflammatory arthritis which occurs more frequently in men than women. In their recent publication examining sex differences in the genetic aetiology of common complex traits and diseases, Bernabeu et al. (2021) observe differences in heritability of AS between sexes, and a genome-wide significant genotype by sex interaction in risk of AS at the major histocompatability (MHC) locus^1^. The authors then present evidence suggesting that this genotype by sex interaction arises primarily as a result of differential expression of the gene *MICA* across the sexes in skeletal muscle tissue. Through a series of conditional association analyses in the UK Biobank, reanalysis of the GTEx gene expression resource and RNASeq experiments on peripheral blood cells from AS cases and controls, we show that the genotype by sex interaction the authors’ report is unlikely to be a result of variation in *MICA*, but probably reflects a known interaction between the *HLA-B* gene, sex and risk of AS. We demonstrate that the diagnostic accuracy of AS in the UK Biobank is low, particularly amongst women, likely explaining some of the observed differences in heritability across the sexes and the difficulty in precisely locating association signals in the cohort.

---

The genetic association between the *B27* allele of the *HLA-B* gene and AS was first reported in the early 1970’s and is one of the strongest known associations between a genetic variant and a common complex disease^2^. Whilst genetic variants at the adjacent *MICA* locus (∼46 kbps from *HLA-B*) have also been reported to be associated with risk of AS^3^, signals at *MICA* dramatically attenuate after conditioning on *HLA-B27*, and likely represent spurious signals that are a consequence of the high linkage disequilibrium across this part of the MHC region^4^. As the frequency of *HLA-B27* is known to differ between male and female AS patients^5^, we were interested in whether *HLA-B27* rather than *MICA* variants might explain the genotype x sex interactions reported in Bernabeu et al (2021)^1^.

We therefore performed a series of conditional logistic regression analyses in the UK Biobank^6^ where we examined the effect of including imputed *HLA-B27* status and its interaction with sex on the results of the model employed in Bernabeu et al (2021)^1^. We found that imputed *HLA-B27* status was more strongly related to risk of AS than rs9266267 (Self-reported AS: p = 2.2×10^−308^ vs p = 2.5×10^−198^), the variant reported by Bernabeu et al as having the strongest evidence for interaction with sex (also this SNP is closer physically to *HLA-B* than *MICA*). Indeed, the signal at rs9266267 was severely attenuated after conditioning on *HLA-B27* status (p = 0.63), whereas the association between AS risk and *HLA-B27* remained strongly significant after conditioning on rs9266267 (p = 3.2×10^−111^). Likewise, inclusion of *HLA-B27* and a *HLA-B27* x sex interaction term in the logistic regression completely obviated evidence for an interaction between rs9266267 and sex (p = 0.8) in determining risk of AS. In contrast, evidence for the *HLA-B27* x sex interaction term (p = 9.1×10^−6^) remained. Inclusion of covariate terms involving *HLA-B27* status also attenuated evidence for interaction between rs9266267 and sex when AS was defined on the basis of in-patient hospital records (Extended Data 1), although the low quality of AS diagnoses in the UKBB (see below) makes localization of signals in this region through conditional analyses challenging.

In their manuscript, Bernabeu et al. (2021) report excluding genetic variants with a MAF < 10% in their analyses of binary traits due to concerns over the small number of cases available^1^. However, since the frequency of the *HLA-B27* allele in the UKBiobank dataset is ∼4%, it is likely that the authors removed SNP variants that tag *HLA-B27* with high sensitivity and specificity, which would also be expected to have a MAF <10%. Indeed, repeating genetic analyses across this region with a less stringent MAF cut-off (MAF < 1%) revealed several markers in the *MICA* and *HLA-B* gene regions that exhibited much stronger evidence of association with AS, and stronger evidence for genotype by sex interaction than the rs9266267 variant. Indeed, conditioning on imputed *HLA-B27* status removed much of the evidence for genotype x sex interaction across this region (Figure 1).

**Figure 1.**
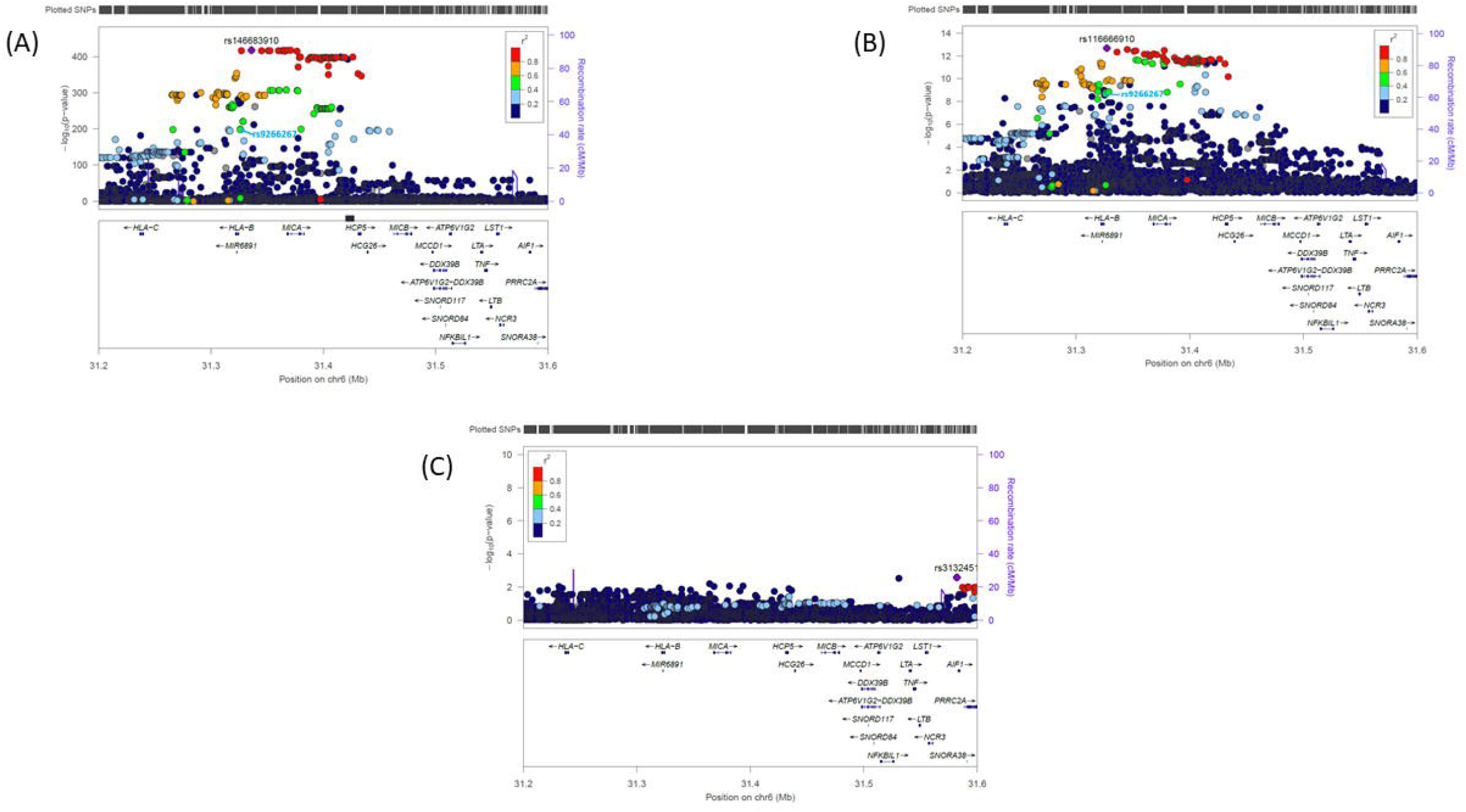
Locus Zoom plots showing minus log_10_ two-sided p-values (y-axis) across the *MICA* and *HLA-B* region of the MHC (x-axis) for (A) the main effect of individual SNPs on AS risk, (B) SNP by sex interaction p-values, and (C) SNP by sex interaction p-values after conditioning on *HLA-B27* status and *HLA-B27* x sex interaction terms. Results for self-reported AS are shown. We note that many SNPs across the region show stronger p-values than those reported for *HLA-B27* status. This is likely a consequence of *HLA-B* imputation error, diagnostic error and also the fact that the relationship between AS risk and *HLA-B27* status is known to be not completely dominant.

Next, we examined the authors’ purported genotype x sex interaction involving the rs56705452 variant and expression of *MICA* in skeletal muscle (reported as p = 1.8×10^−6^ and presented graphically in Figure 8 of the Extended Data in Bernabeu et al. (2021)^1^) using a more recent version of the GTEx dataset. The authors performed these gene expression analyses to investigate a possible mechanism through which the genotype x sex interaction in AS may be mediated. Although such mediation is possible, it would be surprising given that AS is primarily an immunological disease characterized by the involvement of immune, gut epithelial and connective tissues, rather than skeletal muscle. Nevertheless, if genotype x sex interactions involving gene expression data are an uncommon occurrence, then a robust statistical interaction involving the same variant (and of a similar form) could provide circumstantial evidence for the involvement of *MICA* in AS pathology.

Unfortunately, we were unable to replicate the authors’ genotype x sex interaction in the GTEx skeletal muscle data using a more recent version of the resource (i.e. version 8 as opposed to version 6p) which contains a larger number of individuals (p = 0.0013, Figure 2). This magnitude of p-value would not have been considered significant in the original paper, and thus does not provide strong evidence for an interaction between rs56705452, sex and expression of *MICA*. To assess whether MICA expression varied across different immune cell types in AS, we generated two separate RNA-seq datasets^7-10^, one using unsorted peripheral blood mononuclear cells (PBMCs), and the other using PBMCs FACS sorted into CD4, CD8, monocytes, natural killer cells and gamma-delta cells, where RNA-seq was performed separately on each cell type. MICA expression was compared between AS cases and controls for the whole sample (i.e. both sexes together) and for males and females separately. We did not observe significant evidence of differential MICA expression between AS patients and healthy controls in any combination (Extended Data Figure 1, Extended Data Table 1).

**Figure 2.**
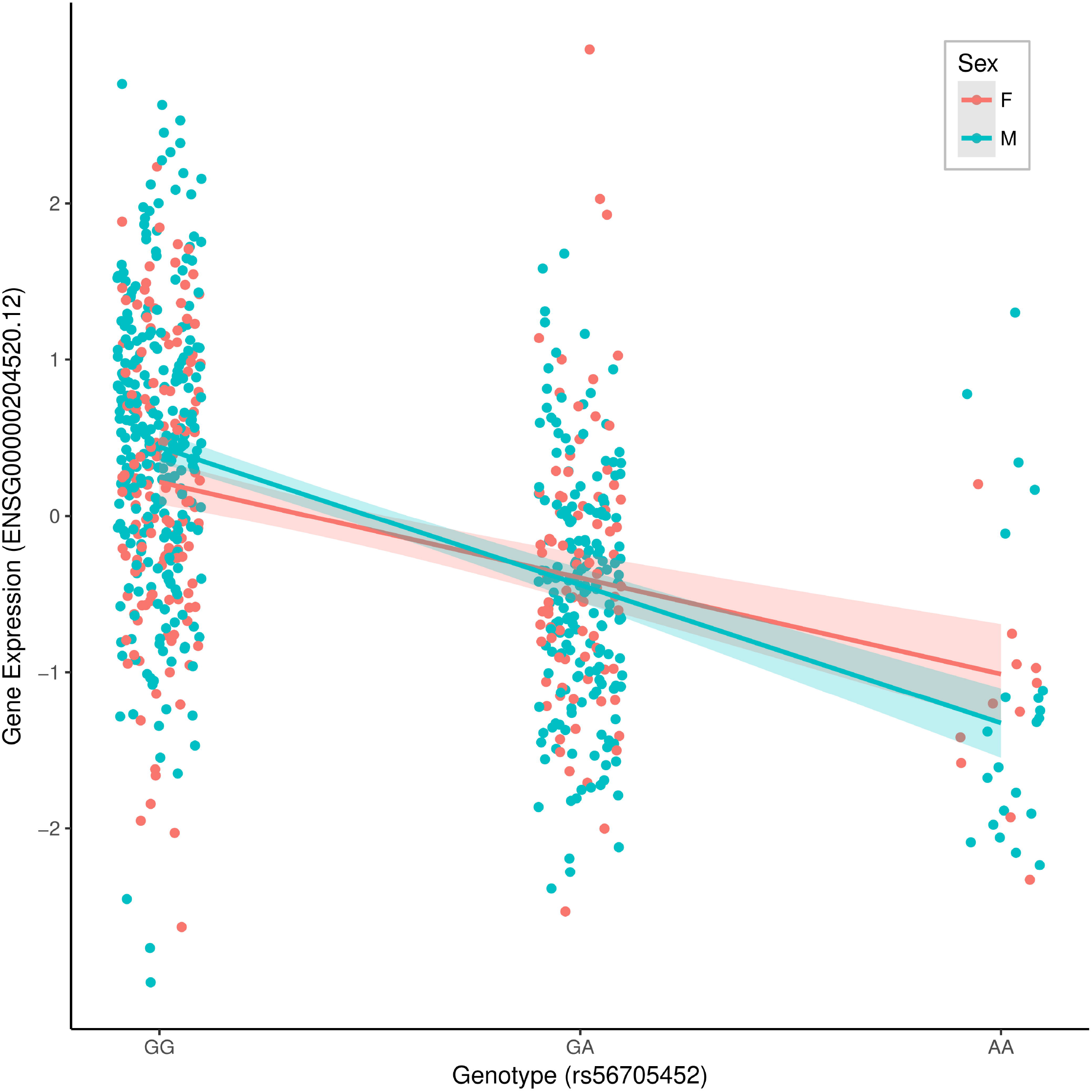
Ordinary least squares linear regression of MICA expression (ENSG00000204520.12) in skeletal muscle tissue (y-axis) on number of copies of the “A” allele at the SNP rs56705452 (x-axis). Individual data points and regression lines with two-sided 95% confidence intervals are shown for males (blue) and females (red).

Finally, diagnosis of AS can be challenging, and experience in biobank datasets shows that misdiagnosis is common, particularly where self-reported^11^. The strong association of *HLA-B27* with the disease (i.e. *HLA-B27* prevalence in clinically diagnosed AS samples is 82-96%^12-14^) enables assessment of diagnostic accuracy of AS in studies. In the UK Biobank, (imputed) *HLA-B27* prevalence amongst AS patients based on hospital inpatient record diagnoses was just ∼68%, and amongst those with a self-reported diagnosis just ∼54%-far lower than the expected figure of 82-96% observed in clinical studies. The substantially lower *HLA-B27* prevalence amongst both the hospital diagnosed and self-reported UK Biobank AS cases, particularly among female cases (Table 1), indicates a high level of misclassification, which would substantially affect heritability analyses such as those reported by Bernabeu et al (2021)^1^.

**Table 1.** *HLA-B27* allele frequency according to diagnostic group and gender. ‘Rheumatologist diagnosed AS’ refers to cases from Li et al. (2021)^14^ where all diagnoses were confirmed by a rheumatologist. UKBB Hospital AS diagnosis data refers to patients with either a primary or secondary hospital record diagnosis of AS (ICD-10 code M45).

| Cohort | Gender | Number | *HLA-B27* allele frequency (%) |
| --- | --- | --- | --- |
| Rheumatologist diagnosed AS | Male | 5724 | 83.8 |
| Rheumatologist diagnosed AS | Female | 2442 | 78.8 |
| Rheumatologist diagnosed AS | Overall | 8166 | 82.3 |
| UKBB Hospital AS diagnosis | Male | 243 | 67.8 |
| UKBB Hospital AS diagnosis | Female | 481 | 46.5 |
| UKBB Hospital AS diagnosis | Overall | 724 | 60.6 |
| UKBB self-reported AS | Male | 844 | 63.3 |
| UKBB self-reported AS | Female | 524 | 38.7 |
| UKBB self-reported AS | Overall | 1368 | 53.9 |

In summary, we have shown through conditional logistic regression analyses that *HLA-B27* is strongly related to risk of AS in the UKBB sample, and that inclusion of terms involving *HLA-B27* status in the regression obviates the differential genotype x sex association between *MICA* variants and risk of AS. We also found little evidence to support a genotype x sex interaction involving the rs56705452 variant and expression of the *MICA* gene in skeletal muscle tissue. Consequently, we contend that known sex differences in *HLA-B27* frequency are likely to explain the genotype x sex interaction observed at the MHC locus in Bernabeu et al. (2021)^1^, and that the association between *MICA* variants and AS reported in the original manuscript is likely to be an artefact brought about by the authors’ exclusion of low frequency variants (MAF <10%) that tag *HLA-B27* status prior to analysis, and the decision not to condition on *HLA-B27* or these same variants. Lastly, the heritability assessments presented, including differences in the strength of genetic contribution to AS in males and females, are likely to have been significantly affected by case misclassification.

## METHODS

### UK Biobank

We performed genetic association analyses on a set of unrelated individuals defined as White British in the UK Biobank (missing rate <2%, total N = 334,996). We ran analyses using two case definitions of AS: (i) self-reported AS (N = 974 cases) and (ii) AS defined by hospital inpatient records (N = 567 cases). For both sets of analyses the remaining white British individuals in the UK Biobank who were not cases in either analysis were used as control individuals (N = 334,752). We performed single locus tests of association across the MHC region ∼1MB either side of the *MICA* gene (i.e. markers on chromosome 6 from 31MB to 32MB Build GRCh37) using logistic regression. We included covariates for sex and the first ten principal components derived from a PCA of the genome-wide SNP data with long regions of linkage disequilibrium including the MHC region removed. We tested for main effects at each SNP across the region, and then subsequently fitted logistic regression models of the form AS status ∼ SNP + sex + SNP x sex, and tested the significance of the SNP by sex interaction term for all markers in the region. Importantly we only excluded variants with MAF <1% from analyses (in contrast to Bernabeu et al. who excluded variants with MAF < 10%).

In order to investigate the robustness of the *MICA*-sex interaction in the UK Biobank, we then performed a series of conditional logistic regression analyses using the same baseline covariates as above (i.e. sex and ten principal components). We fit the following conditional logistic regression models where AS status was the outcome, and the following terms were the predictor variables: (1) the rs9266267 variant (i.e. the *MICA* variant with the strongest evidence for genotype x sex interaction reported in Bernabeu et al. 2021); (2) *HLA-B27* (fitted as a binary term indicating its presence or absence); (3) rs9266267 plus an rs9266267 by sex interaction term; (4) *HLA-B27* plus an *HLA-B27* by sex interaction term; (5) rs9266267, *HLA-B27*, rs9266267 by sex interaction, and *HLA-B27* by sex interaction terms. *HLA-B* alleles were imputed using the HLA*IMP:02 program^15^. Participants with posterior probability of imputed HLA-B27 alleles greater than 0.7 were considered HLA-B27 positive while the samples with posterior probability equal to zero were considered HLA-B27 negative (see Extended Data Table 2 for a breakdown of the 1423 indiduals excluded from analyses because of low quality HLA imputation). To ensure comparability, all logistic regression analyses were performed on samples for which *HLA-B27* status could be assigned with confidence. All logistic regression analyses were performed using the R software package, and significance was determined via Wald tests with two tailed p-values.

### Clinically defined AS set

MICA expression was analysed in two separate RNA-seq datasets. AS cases and controls were recruited, and RNA-sequencing performed, as previously reported^7^. The first dataset consisted of 144 PMBC samples from 66 AS patients (56 male and 10 female) and 78 healthy controls (45 male and 33 female). The second dataset contained 100 individuals with 51 AS patients (39 male and 12 female) and 49 healthy controls (20 males and 29 females). Prior to sequencing the second dataset, cells were FACS sorted on a MoFlo Astrios (Beckman Coulter) into CD4, CD8, monocytes, natural killer cells and gamma-delta cells, and purity of isolated cells was determined using FlowJo software (BD Biosciences). RNA-seq was performed separately on each cell type. Transcripts were quantified with Salmon (v1.5.1)^9^ and converted to gene-level counts for analysis with DESeq2^8^ via tximport^10^. Each dataset was analysed as a whole and separately for males and females. Differences in expression levels between cases and controls were assessed using negative binomial regression. Ethics approval for collection of blood samples was granted by the Princess Alexandra Hospital and the Queensland University of Technology (QUT) Ethics Committees (Metro South approval no. HREC/05/QPAH/221 and QUT approval no. 1600000162).

### GTEx analyses

We examined evidence for interaction between rs56705452, sex and expression levels of *MICA* (ENSG00000204520.12) in skeletal muscle tissue using the v8 release of GTEx (469 males and 237 females). Following the GTEx Consortium eQTL discovery pipeline, we fit an ordinary least squares linear regression to test the effect of the rs56705452 by sex interaction with expression of *MICA* as the outcome and rs56705452, sex, five genetic principal components, 60 PEER factors, and genotyping platform and protocol terms as covariates. To evaluate significance of terms, t tests with two tailed p-values were computed.

## Data Availability

This research has been conducted using the UK Biobank resource (References 53641 and 21024). We thank the participants of UK Biobank for making this work possible. The UK Biobank genotype and phenotype data are available by application from https://www.ukbiobank.ac.uk/. GTEx data are available upon application through dbGAP as described at https://gtexportal.org/home/datasets.

## Acknowledgements

This research has been conducted using the UK Biobank resource (References 53641 and 21024). D.M.E. is funded by an Australian National Health and Medical Research Council Senior Research Fellowship (APP1137714). A.F.M is funded by an Australian Research Council Future Fellowship (FT200100837). G.W. is supported by the University of Queensland Graduate School Scholarship (UQGSS). Z.L. is funded by Queensland University of Technology Vice-chancellor Reseach Fellowship.

## AUTHOR INFORMATION

### Contributions

Z.L., A.F.M., G.W. and J.J.E. performed data analysis. J.W. and T.J.K. performed FACS of blood samples. J.W. and T.J.K. assisted in sample collection. J.W. assisted with RNASeq of ankylosing spondylitis samples. Sample collection and RNASeq of ankylosing spondylitis samples was led by M.A.B. M.A.B. and D.M.E. wrote the manuscript. All authors reviewed the manuscript.

### Corresponding Author

Correspondence to David Evans.

## ETHICS DECLARATION

### Competing Interests

The authors declare no competing interests.

**Extended Data 1**

When comparing individuals with AS defined from in-patient hospital records, *HLA-B27* was more strongly related to risk of AS than rs9266267 (p = 7.7× 10^−247^ vs p = 3.3×10^−152^). Indeed, the signal at rs9266267 was severely attenuated after conditioning on *HLA-B27* status (p = 0.014), whereas the association between AS risk and *HLA-B27* remained strongly significant after conditioning on rs9266267 was (p = 5.6×10^−73^). Inclusion of *HLA-B27* and a *HLA-B27* x sex interaction term in the logistic regression substantially reduced evidence for an interaction between rs9266267 and sex (p = 0.38).

**Extended Data Figure 1.**
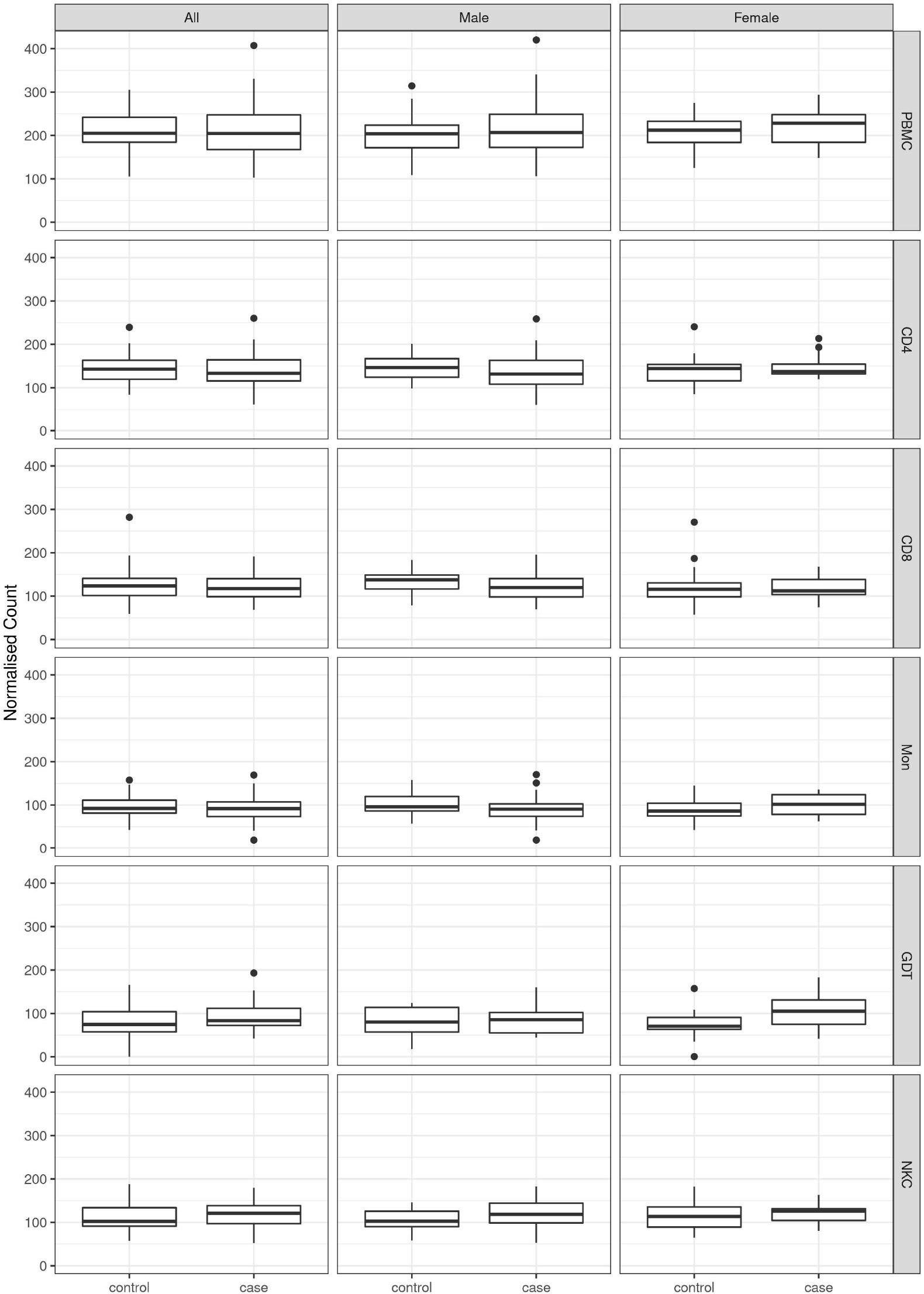
Boxplots showing the distribution of normalised RNA-seq counts between AS cases and healthy controls for the PBMC data set and FACS sorted single cell type datasets (CD4, CD8, Monocytes, gamma-delta cell and natural killer cells). Normalised counts were obtained from DESeq2^8^. Boxes represent the median, lower and upper quartiles of count data. Whiskers extend 1.5 times the interquartile range in both directions. (PBMC = Peripheral blood mononuclear cells; CD4 = CD4+ T cells, CD8 = CD8+ T cells, Mon = Monocytes, GDT = gamma-delta T cells; NKC = natural killer cells).

**Extended Data Table 2.** The breakdown numbers of 1,423 UKBB participants whose *HLA-B27* status was classified as unknown.

| Cohort | Gender | Number |
| --- | --- | --- |
| UKBB Hospital AS diagnosis | Male | 9 |
| UKBB Hospital AS diagnosis | Female | 2 |
| UKBB Hospital AS diagnosis | Overall | 11 |
| UKBB self-reported AS | Male | 10 |
| UKBB self-reported AS | Female | 4 |
| UKBB self-reported AS | Overall | 14 |
| UKBB AS | Male | 12 |
| UKBB AS | Female | 5 |
| UKBB AS | Overall | 17 |
| UKBB controls | Male | 617 |
| UKBB controls | Female | 789 |
| UKBB controls | Overall | 1406 |

