## Extended_Data_Table1 for "*HLA-B27* and not variation in *MICA* is responsible for genotype by sex interaction in risk of Ankylosing Spondylitis"

**Extended Data Table 1.** Expression of MICA in PBMCs

| Sample | Base Mean | log2FoldChange | lfcSE | stat | pvalue |
|---|---|---|---|---|---|
| PBMC_All | 210.2893657 | 0.043252883 | 0.053749 | 0.804726 | 0.420978 |
| PBMC_Male | 209.0874955 | 0.062139702 | 0.065172 | 0.953472 | 0.340351 |
| PBMC_Female | 212.0897046 | 0.019896796 | 0.110558 | 0.179967 | 0.857179 |
| CD4_All | 140.8674817 | -0.032716973 | 0.072073 | -0.45394 | 0.649872 |
| CD4_Male | 139.6674791 | -0.114843903 | 0.106838 | -1.07494 | 0.282402 |
| CD4_Female | 142.3323067 | 0.090802969 | 0.111296 | 0.815866 | 0.414577 |
| CD8_All | 122.330594 | -0.082791202 | 0.078304 | -1.05731 | 0.290371 |
| CD8_Male | 124.414853 | -0.140904966 | 0.099877 | -1.41079 | 0.158307 |
| CD8_Female | 119.0159064 | -0.010181828 | 0.152312 | -0.06685 | 0.946702 |
| Mon_All | 92.99157838 | -0.039975483 | 0.086441 | -0.46246 | 0.64375 |
| Mon_Male | 93.481456 | -0.185249817 | 0.121519 | -1.52445 | 0.127397 |
| Mon_Female | 92.05611043 | 0.182257338 | 0.139917 | 1.302608 | 0.192708 |
| GDT_All | 84.52028438 | 0.224546173 | 0.171878 | 1.306424 | 0.191408 |
| GDT_Male | 83.88559349 | 0.063120715 | 0.250968 | 0.251509 | 0.801421 |
| GDT_Female | 84.59075764 | 0.454842134 | 0.241668 | 1.882094 | 0.059823 |
| NKC_All | 113.8967404 | 0.107705677 | 0.083043 | 1.296992 | 0.194634 |
| NKC_Male | 113.9205611 | 0.175065421 | 0.110365 | 1.586245 | 0.112684 |
| NKC_Female | 113.5662037 | 0.083779329 | 0.152986 | 0.547629 | 0.583947 |

PBMC = Peripheral blood mononuclear cells; CD4 = CD4+ T cells, CD8 = CD8+ T cells, Mon = Monocytes

Base Mean = Base mean expression across all samples

log2FoldChange = log2 fold change control vs AS

lfcSE = Standard error of the log2FoldChange

stat = Wald statistic

pvalue = P-value

padj = Adjusted P-value FDR

| padj |
|---|
| 0.652214 |
| 0.636481 |
| 0.968416 |
| 0.89346 |
| 0.698834 |
| 0.842222 |
| 0.834939 |
| 0.631376 |
| 0.999961 |
| 0.838504 |
| 0.562952 |
| 0.617102 |
| 0.86626 |
| 0.972364 |
| 0.997506 |
| 0.691064 |
| 0.578698 |
| 0.999255 |

;, GDT = gamma-delta T cells; NKC = natural killer cells
